# Evaluating the Impact of Diagnostic Stewardship in Community-Acquired Pneumonia with Syndromic Molecular Testing: A Randomized Clinical Trial

**DOI:** 10.1101/2023.11.11.23298408

**Authors:** Dagfinn L. Markussen, Sondre Serigstad, Christian Ritz, Siri T Knoop, Marit H Ebbesen, Daniel Faurholt-Jepsen, Lars Heggelund, C. H. Henri van Werkhoven, Tristan W Clark, Rune O Bjørneklett, Øyvind Kommedal, Elling Ulvestad, Harleen M.S. Grewal

## Abstract

**Importance:** Lower respiratory tract infections, including community-acquired pneumonia (CAP), are a leading cause of hospital admissions and mortality. An aetiological diagnosis of CAP is delayed due to long turnaround times with laboratory testing. Rapid microbiologic diagnosis is imperative for the management of CAP and may limit antibiotic overuse. Molecular tests have the potential to optimize treatment decisions and management of CAP, but limited evidence exists to support their routine use.

**Objective:** To determine whether the use of a syndromic PCR-based panel for rapid testing of CAP in the emergency department (ED) leads to faster, more accurate microbiology-result-based treatment.

**Design, Setting, and Participants:** A pragmatic, parallel-arm, single-blinded, single-centre, randomised controlled superiority trial conducted in the emergency department of a large tertiary care Norwegian hospital, where adult patients with suspected CAP were recruited.

**Intervention:** Patients were randomly assigned 1:1 to rapid syndromic molecular panel testing (FAP-plus) of lower respiratory tract (LRT) samples and standard-of-care or standard-of-care alone.

**Main Outcomes and Measures:** Primary outcomes were the provision of pathogen-directed treatment based on a microbiological test result and the time to provision of pathogen-directed treatment (within 48h from randomisation).

**Results:** Between Sep 25, 2020, and Jun 21, 2022, 374 patients were enrolled, with 187 in each arm. Analysis of primary outcomes showed that 66 (35%) of 187 patients in the FAP-plus arm and 24 (13%) of 187 patients in the standard-of-care arm received pathogen-directed treatment corresponding to a reduction in absolute risk of 21.9% (95% CI 13.5–30.3%) and an OR for the FAP-plus arm of 3.53 (95% CI 2.13–6.02; p<0.0001). The mean time to provision of pathogen-directed treatment within 48h was 34.5h in the FAP-plus arm and 43.8h in the standard-of-care arm (mean difference -9.4h, 95% CI -12.7– -6.0h; p<0.0001). The corresponding hazard ratio for FAP-plus compared to standard of care was 3.08 (95% CI 1.95–4.89). Findings remained unaltered after adjustment for season.

**Conclusions and Relevance:** The routine deployment of PCR testing for LRT-pathogens enables faster and more targeted microbial treatment for patients with suspected CAP. Rapid molecular testing could complement or replace selected standard time-consuming laboratory-based diagnostics.

**Trial registration:** ClinicalTrials.gov Identifier: NCT04660084

**Key Points:** *Question:* Does the judicious use of a syndromic PCR-based panel for rapid testing of patients hospitalised with suspected community-acquired pneumonia (CAP) lead to faster, more accurate microbiology-result-based treatment?

*Findings:* In this randomised controlled diagnostic stewardship trial, molecular testing significantly in-creased the proportion of suspected CAP patients that received pathogen-directed treatment and reduced the mean time to pathogen-directed treatment by 9.4h compared to standard-of-care. A syndromic PCR-based result was delivered within 4 hours for all CAP patients.

*Meaning:* The routine deployment of PCR testing for LRT-pathogens enables faster and targeted microbial treatment for patients with suspected CAP. Rapid molecular testing could complement or replace selected standard time-consuming laboratory-based diagnostics.

## Introduction

Lower respiratory tract (LRT) infections, including community-acquired pneumonia (CAP), are a leading cause of hospital admissions and mortality.^1–3^ Despite their significant impact, most patients do not receive a microbiological diagnosis and targeted treatment.^4^ Moreover, the current gold standard for bacteriological diagnosis remains culture-based methods that are labour-intensive, only detect a pathogen in 20-40% of patients and are insufficient to influence early decisions on antimicrobial therapy^4,5^.

Rapid syndromic PCR-based panels have improved pathogen detection, potentially facilitating pathogen-directed treatment, reducing unnecessary use of antibiotics, and shortening length of hospital stay (LOS).^6,7^ Although the potential benefits of rapid PCR panels in CAP are clear, limited evidence currently supports their routine use. A few trials have examined patients with respiratory tract infections (RTIs) using molecular point-of-care tests (mPOCT) for a combination of viruses and atypical bacteria with modest and conflicting results on antibiotic use and LOS.^8–11^ A recent randomised trial examined pneumonia patients with a comprehensive mPOCT, finding an increase in results-directed therapy and de-escalation of antibiotics in the mPOCT group.^12^ However, the study included a mixture of patients from intensive care units with hospital-acquired-ventilator-associated- and community-acquired pneumonia, findings that may not be specifically applicable to CAP patients.

In this study, we aim to establish whether the judicious use of a syndromic PCR-based panel for rapid testing of CAP in the emergency department (ED) leads to faster, more accurate microbiology-result-based treatment. The results of this trial have the potential to inform future guidelines for the management of CAP.

## Methods

### Study design

This study was a pragmatic, parallel-arm, single-blinded, single-centre, randomised controlled superiority trial conducted in a large Norwegian hospital (Haukeland University Hospital) that serves as a local hospital for a population of approximately 470,000 and a referral hospital for 1,000,000 inhabitants. The study protocol^13^ was approved by the Regional Committee for Medical and Health Research Ethics, Norway (registration no. 31935). ClinicalTrials.gov NCT04660084.

### Patients

Patients were eligible for inclusion if they were ≥18 years of age and presented to the ED, with suspicion of CAP and fulfilled at least two of the following criteria: new or worsening cough; new or worsening expectoration; new or worsening dyspnoea; haemoptysis; pleuritic chest pain; radiological evidence of pneumonia; abnormalities on chest auscultation and/or percussion; fever (≥38.0⁰C). Patients were ineligible if they had cystic fibrosis, severe bronchiectasis, hospitalization within the last 14 days prior to admission, a palliative approach was being taken (defined as life expectancy below two weeks), or if they were not willing to provide an LRT sample. ^13^ Written informed consent was obtained from all patients or from their legal guardian/close relatives.

### Randomisation and masking

Patients were randomly assigned 1:1 to receive syndromic PCR (FAP-plus arm) testing in addition to standard-of-care microbiology diagnostics or to receive standard-of-care alone. Block randomisation with randomly varying block size (4, 6, or 8 patients per block) was applied using the **R** package “blockrand”. The allocation sequence was implemented in the electronic data capture system Viedoc (Viedoc Technologies, Sweden). Trial participants and clinical care providers in the ED were blinded to group allocation.

### Procedures

Eligible patients were included shortly after presenting to the ED. Study nurses/investigating physicians collected baseline information.

Rapid testing was performed using the BioFire FilmArray Pneumonia panel *plus* (FAP-plus) (bioMérieux, France). The FAP-plus PCR panel detects 27 bacterial and viral respiratory pathogens with seven antimicrobial resistance genes. Standard-of-care methods included: blood cultures, the pneumococcal urine test (Quidel Corporation, U.S.), and an in-house PCR for oro- and/or nasopharyngeal swabs targeting respiratory viruses and atypical bacteria (influenza A and B viruses, human parainfluenza viruses 1-3, respiratory syncytial virus (RSV), human metapneumovirus, rhinovirus, coronavirus (229E, OC43, HKU1, NL63), *Bordetella pertussis*, *Bordetella parapertussis*, *Mycoplasma pneumoniae* and *Chlamydophilia pneumoniae*.

Results for the FAP-plus test, blood cultures, and a positive urine test for *S. pneumoniae* or *Legionella pneumophilia*, were communicated telephonically to the treating staff for both arms. The phone call alerted staff that a test result was available in the patient’s electronic medical journal, while the report in the journal provided results, including standard responses suggesting whether the detected bacteria was a likely pathogen (eTable 1 in Supplement 2).

At admission, all patients were tested for SARS-CoV-2 using oro-/nasopharyngeal swabs tested on the GeneXpert system (Cepheid, U.S.). LRT-samples were collected in the ED by sputum induction using nebulized isotonic (0.9%) or hypertonic (5.8%) saline. Endotracheal aspiration was performed in case of unsuccessful sputum induction and in patients with SARS-CoV-2.

### Outcomes

Two primary outcomes were defined: 1) provision of pathogen-directed treatment based on a relevant microbiological test result and 2) time to the provision of pathogen-directed treatment (within 48h from randomisation). The former is a binary outcome, whereas the latter is an event-time outcome where right censoring is present, i.e., patients may cease to be in the study due to death, discharge, or reaching 48h (from the time of randomisation) without having received pathogen-directed treatment. Primary outcomes were defined for all patients that were randomised.

Two study physicians assessed whether (and when) a patient received pathogen-directed antimicrobial treatment based on a microbiological result. In case of disagreement, a third physician arbitrated. To be considered as pathogen-directed treatment, documentation in the patient’s journal by the treating physician(s) was required, describing (a) change in antimicrobial treatment, or (b) continuation of already correctly initiated antimicrobial treatment, or (c) discontinuation of antimicrobial treatment. The final diagnosis of CAP was established retrospectively through clinical adjudication using pre-specified criteria (eTable 2 in Supplement 2).

Secondary outcomes included the binary outcomes: provision of any antibiotics, provision of narrow-spectrum antibiotics within 48h, provision of a single dose of antibiotics, antibiotics not used for more than 48h, treatment with intravenous antibiotics, de-escalation from broad-spectrum to narrow-spectrum antibiotics, and escalation from narrow-spectrum to broad-spectrum antibiotics. Continuous secondary outcomes included the duration of provision of antibiotics (days), intravenous antibiotics, broad-spectrum antibiotics, time elapsed from admission to the administration of antibiotics, and turnaround time defined as the time from admission to receiving a microbiological report (FAP-plus result and/or sputum culture) (in hours). Broad-spectrum antibiotics were defined as penicillin with enzyme inhibitors, second and third-generation cephalosporins, carbapenems and quinolones.^14^ LOS in days, mortality (30 days and 90 days), readmission within 30 days after discharge, and adverse outcomes were also reported.

### Statistical analysis

As two primary outcomes were used, separate sample size calculations were performed for each outcome at a two-sided significance level of 0.05/2 = 0.025 (instead of 0.05), assuming a power of 80%. To detect an increase in the provision of pathogen-directed treatment from 0.40 to 0.50, the required sample size was 470 per arm. Likewise, to detect a reduction of 0.2 standard deviation in the time to provision of pathogen-directed treatment, the sample size was found to be 477 per arm, i.e., in total, 954. Allowing for a 10% dropout rate resulted in a total sample size of 1060 patients.

Baseline patient characteristics were summarized using percentage, count, and total for categorical variables and median and interquartile range (IQR) for continuous variables. Missing baseline values were imputed by means of a single imputation using chained equations.^15^

The two primary outcomes were analysed according to the intention-to-treat principle. Available-case analyses were used for the secondary outcomes.

For binary outcomes, logistic regression models with logit and identity link functions were used to estimate odds ratios and absolute risk differences, respectively. Two models were fitted for the event-time primary outcome: Cox regression and a restricted mean survival time model.^16^ The proportional hazards assumption for Cox regression was assessed visually using cumulative log-log plots. The restricted mean survival time model does not require proportional hazards.^17^ Kaplan-Meier and log-rank tests were also reported. For continuous secondary outcomes, linear regression with logarithm-transformed outcomes was used to estimate differences in medians and ratios of medians; differences in medians were approximated using a Taylor expansion.^18^ For the two primary outcomes, analyses adjusted for season was also carried out. A significance level of 0.05 was applied and the statistical software **R** was used (version 4.1.2).^19^ Imputation through chained equations and fitting of restricted mean survival time models were carried out using the **R** packages mice and SURVRM2, respectively.

## Results

Patients were recruited from Sep 25, 2020, to Jun 1, 2021, and again from Aug 15, 2021, to Jun 21, 2022. The trial was stopped earlier than planned as an ad hoc interim analysis conducted due to slow recruitment, carried out Jun 16, 2022, showed highly significant differences between the two arms for both primary outcomes. A total of 2265 patients were assessed for eligibility whereof 374 patients were included, with 187 patients randomized to each arm (figure 1). Among patients randomised and providing a sputum sample, 200 had a diagnosis of CAP; 97 in the FAP-*plus* arm and 103 in the standard-of-care arm.

**Figure 1:**
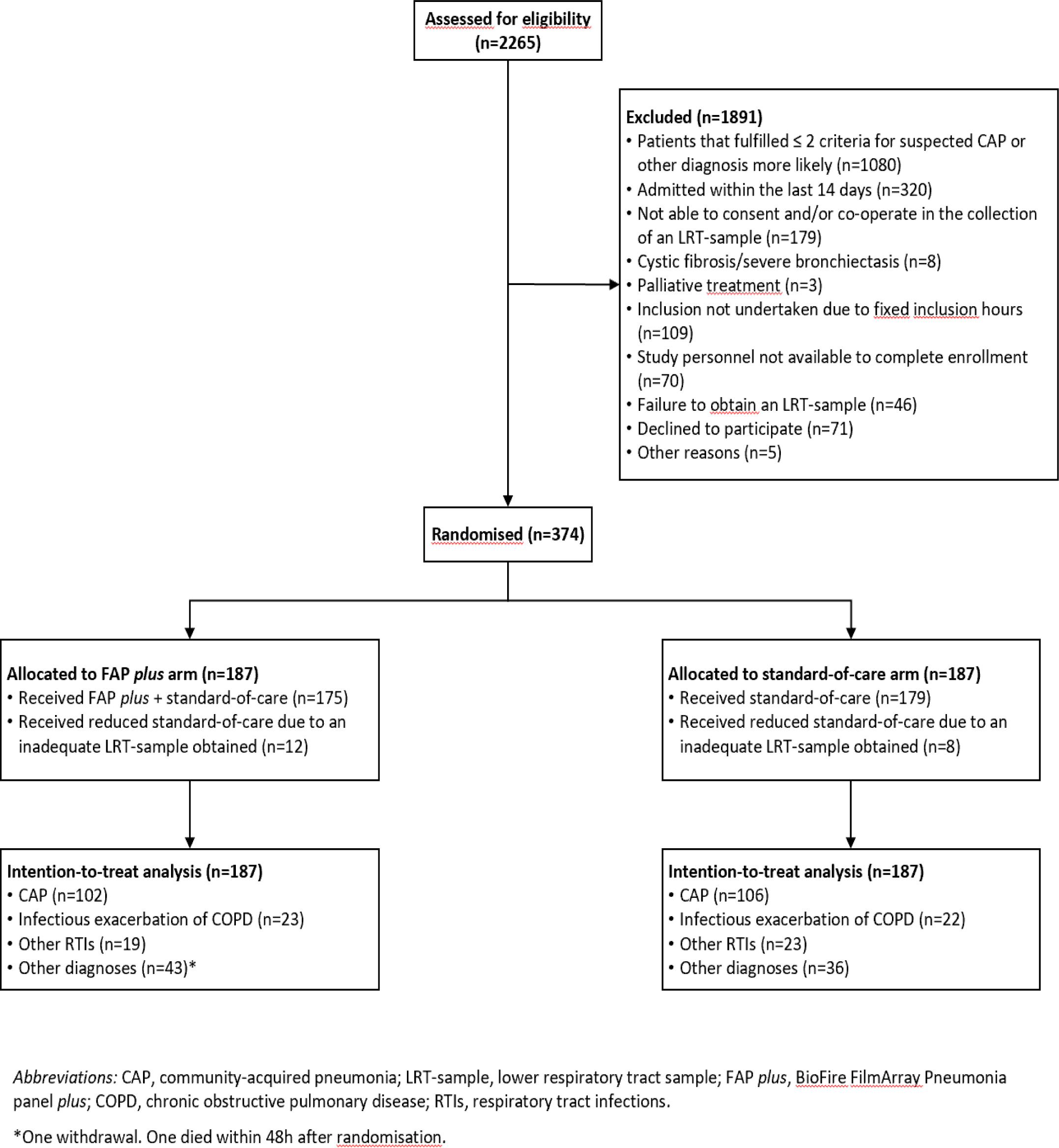
study profile.

### Baseline characteristics

Randomisation produced similar distributions of patient characteristics in both arms (Table 1). The median age of patients was 72 years (IQR 60–79 years) and 153 (41%) of 374 participants were female. Distributions were also comparable between arms for the subgroup of CAP patients (eTable 3 in Supplement 2). CAP patients had characteristics that were similar to all patients randomised with the exception of CRP, where the median level was 93 mg/L (IQR 38–178 mg/L) among all patients and 145 mg/L (IQR 89–219 mg/L) among CAP patients.

**Table 1:** Baseline characteristics for all randomised patients (n=374)

|  | **FAP-plus**<br>(n=187) | **Standard-of-care**<br>(n=187) |
| --- | --- | --- |
| *Age (years)* | 73 (61, 79) | 71 (60, 80) |
| *Sex (% Female)* | 39 (73/187) | 43 (80/187) |
| *Current smoker* | 20 (37/187) | 19 (36/187) |
| *Annual influenza vaccine* | 52 (97/187) | 52 (98/187) |
| *Pneumococcal vaccine in the last five years* | 40 (74/187) | 39 (73/187) |
| *Duration of symptoms prior to admission (days)* | 6.6 (3.6, 10.5) | 5.7 (3.5, 9.5) |
| *Antibiotics within 48 hours* | 3 (6/187) | 1 (2/187) |
| *Antibiotics within the last month (%)* | 17 (32/155) | 11 (20/167) |
| **Comorbidities** |  |  |
| *Hypertension* | 41 (76/187) | 37 (70/187) |
| *Cardiovascular disease* | 31 (58/187) | 34 (64/187) |
| *Respiratory disease* | 55 (102/187) | 55 (103/187) |
| *Renal disease* | 12 (23/187) | 9 (16/187) |
| *Liver disease* | 0 (0/187) | 1 (2/187) |
| *Diabetes* | 13 (25/187) | 16 (29/187) |
| *Immunocompromised* | 7 (14/187) | 11 (20/187) |
| *Cancer* | 11 (20/187) | 6 (11/187) |
| *Charlson Comorbidity Index score* | 4 (2, 5) | 4 (2, 5) |
| **Observations** |  |  |
| *Temperature (°C)* | 37.1 (36.7, 37.5) | 37.0 (36.8, 37.5) |
| *Pulse rate (bpm)* | 94 (79, 108) | 93 (83, 105) |
| *Respiratory rate (bpm)* | 22 (20, 26) | 24 (20, 28) |
| *O<sub>2</sub> saturation (%)* | 93 (90, 96) | 94 (89, 97) |
| *Supplementary O<sub>2</sub>* | 8 (15/187) | 6 (11/187) |
| *Blood pressure (mmHg)* |  |  |
| *Systolic* | 131 (117, 150) | 135 (118, 149) |
| *Diastolic* | 80 (71, 89) | 81 (70, 90) |
| ***Laboratory and radiology*** |  |  |
| *CRP (mg/L)* | 81 (31, 159) | 108 (47, 199) |
| *White blood cell count*<br>*($\times 10^9$ per L)* | 10.5 (8.1, 15.1) | 10.7 (8.1, 13.8) |
| *Chest X-ray (%)* | 96 (180/187) | 93 (174/187) |
| *Chest CT (%)* | 26 (48/187) | 26 (49/187) |
| *CURB-65* | 1 (1, 2) | 1 (1, 2) |
| *qSOFA* | 1 (0, 1) | 1 (0, 1) |
| *SARS-CoV-2 positive test* | 15 (28/187) | 13 (24/187) |

Data are shown as percentage (count) and median (IQR) for binary and continuous variables, respectively. Missing data (FAP-plus: n=10, Standard-of-care: n=7) were imputed using multiple imputations through chained equations.
FAP-plus, BioFire FilmArray Pneumonia panel *plus* (bioMérieux S.A., Marcy-l'Etoile, France); IQR, interquartile range; O<sub>2</sub>, oxygen; CRP, C-reactive protein; CT, computed tomography; CURB-65, confusion, blood urea nitrogen, respiratory rate, blood pressure and age > 65 years; qSOFA, quick sequential organ failure assessment score. CRP and white blood cell counts presented are the highest values during hospitalization.

### Findings for all randomised patients

Forty-eight hours from randomisation, 66 of 187 (35%) patients in the FAP-plus arm and 25 of 187 (13%) patients in the standard-of-care arm received pathogen-directed treatment (figure 2), corresponding to a reduction in absolute risk of 21.9% (95% CI 13.5–30.3%) and an OR for the FAP-*plus* arm of 3.53 (95% CI 2.13–6.02; p<0.0001; Table 2).

**Figure 2:**
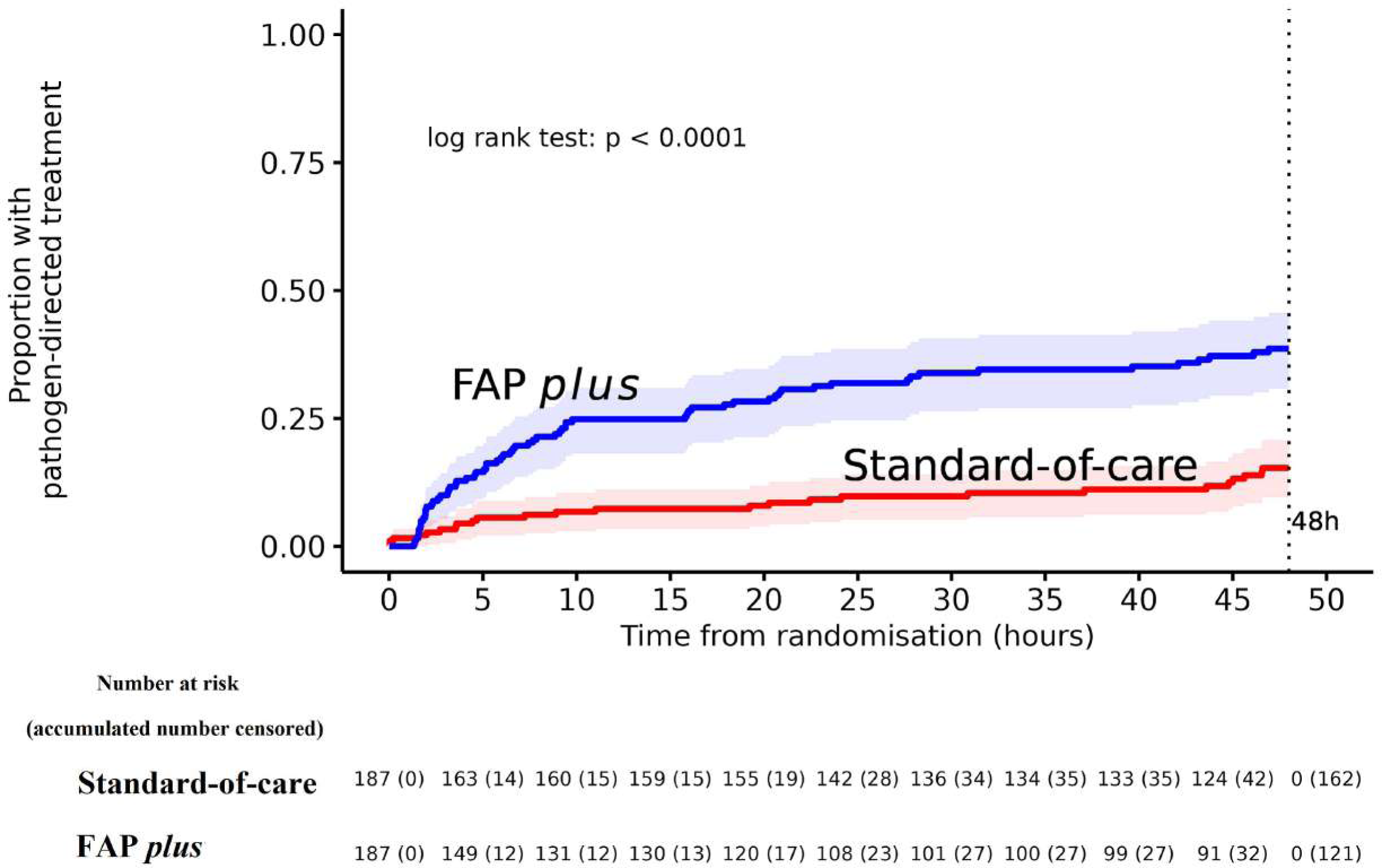
Proportion of patients receiving pathogen-directed treatment (intention-to-treat analysis set) FAP *phs*: BioFire FilmArray Pneumonia panel plus (bioMérieux S.A., Marcy-l’Etoile, France)

**Table 2:** Comparison of rapid testing by FAP-plus and standard-of-care for all randomised patients (n=374)

|  | **FAP-plus**<br>**(n=187)** | **Standard-**<br>**of-care**<br>**(n=187)** | **FAP-plus vs. Standard-of-care<sup>#</sup>** |  |  |  |  |  |
| --- | --- | --- | --- | --- | --- | --- | --- | --- |
| **Outcomes** |  |  | **Unadjusted** |  |  | **Adjusted for season (2020 vs.**<br>**2021)** |  |  |
|  |  |  | **Difference** | **Odds ratio** | **p-value** | **Difference** | **Odds ratio** | **p-value** |
| **Primary**<br>**outcomes** |  |  |  |  |  |  |  |  |
| *Provision of*<br>*pathogen-*<br>*directed*<br>*treatment (%)<sup>§</sup>* | 35.3<br>(66/187) | 13.4<br>(25/187) | 21.9<br>(13.5, 30.3) | 3.53<br>(2.13, 6.02) | <0.0001 | 22.1<br>(13.8, 30.5) | 3.54 (2.13,<br>6.02) | <0.0001 |
| *Provision of any*<br>*antibiotics (%)* | 85.0<br>(159/187) | 84.0<br>(157/187) | 1.1<br>(-6.3, 8.4) | 1.09<br>(0.62, 1.91) | 0.78 | 0.9<br>(-6.4, 8.1) | 1.09<br>(0.62, 1.91) | 0.77 |
|  |  |  |  | **Hazard**<br>**ratio** |  |  | **Hazard**<br>**ratio** |  |
| *Time to*<br>*provision of*<br>*pathogen-*<br>*directed*<br>*treatment (h)<sup>§</sup>* | 34.5<br>(31.6,<br>37.3) | 43.8<br>(42.0,<br>45.6) | -9.4<br>(-12.7, -6.0) | 3.08<br>(1.95, 4.89) | <0.0001 | -9.4<br>(-12.7, -6.0) | 3.08<br>(1.95, 4.89) | <0.0001 |

Data shown as percentage (count) binary outcomes and restricted mean and 95% confidence interval for the event-time outcome, respectively.
<sup>#</sup>: Differences were estimated as risk differences for binary outcomes (on use) and mean differences for event-time outcomes using logistic regression (with the identity link function) and restricted mean survival time methods, respectively. Odds ratios for binary outcomes (on use) and hazard ratios for event-time outcomes were estimated using logistic regression and Cox regression, respectively. p-values corresponding to testing ratios equal to 1 were reported.
<sup>§</sup>: Time elapsed from randomisation to provision of pathogen-directed treatment and within 48h.
FAP-plus, BioFire FilmArray Pneumonia panel *plus* (bioMérieux S.A., Marcy-l'Etoile, France); CAP, community-acquired pneumonia; IQR, interquartile range.

The mean time to provision of pathogen-directed treatment within 48h was 34.5h in the FAP-plus arm and 43.8h in the standard-of-care arm (mean difference -9.4h, 95% CI -12.7– -6.0h; p<0.0001). The corresponding hazard ratio for FAP-plus compared to standard of care was 3.08 (95% CI 1.95–4.89). Findings remained unaltered after adjustment for season.

### Findings for CAP patients

In the FAP-*plus* arm, 46 of 97 (47%) CAP patients and 16 of 103 (16%) patients in the standard-of-care arm received pathogen-directed treatment within 48h (absolute risk difference 30.9%, 95% CI 18.7–43.2%; p<0.0001; table 3), corresponding to an OR for FAP-plus of 4.56 (95% CI 2.41–8.98; p<0.0001). Pathogen-directed treatment within 48h resulted in an escalation to more broad-spectrum antimicrobial treatment in 14 of 97 (14%) and 4 of 103 (4%) patients in the FAP-plus and standard-of-care arms, respectively (absolute difference 10.5%, 95% CI 2.6, 18.5%; p=0.009). Likewise, empirical antibiotic treatment was deescalated to more narrow-spectrum treatment for 10 of 97 (10.3%) and 5 of 103 (4.9%) patients in the FAP-plus and standard-of-care arms, respectively (absolute difference 5.5%, 95% CI -1.9, 12.8%; p=0.14). Further details for the comparison of rapid testing using FAP-plus and standard-of-care for CAP patients, are presented in Table 3. A breakdown of pathogen-directed treatment within 48 hours for CAP patients is provided in eTable 4 in Supplement 2.

**Table 3:** Comparison of rapid testing by FAP-plus and standard-of-care for CAP patients only (n=200)

|  | FAP-plus<br>(n=97) | Standard-of-care<br>(n=103) | FAP-plus vs. Standard-of-care<sup>#</sup> |  |  |
| --- | --- | --- | --- | --- | --- |
| **Outcomes on provision**<br>**(%)** |  |  | **Difference** | **Odds ratio** | **p-value** |
| *Any antibiotics* | 95.9 (93/97) | 95.1 (98/103) | 0.7 (-5.0, 6.5) | 1.19 (0.30, 4.92) | 0.80 |
| *Pathogen-directed treatment (%)<sup>§</sup>* | 47.4 (46/97) | 15.5 (16/103) | 31.9 (19.7, 44.0) | 4.83 (2.52, 9.68) | <0.0001 |
| - Continuation of appropriate empirical treatment | 16.5 (16/97) | 6.8 (7/103) | 9.7 (0.9, 18.5) | 2.66 (1.07, 7.33) | 0.03 |
| - Escalation to more broad-spectrum treatment | 14.4 (14/97) | 3.9 (4/103) | 10.5 (2.6, 18.5) | 4.04 (1.37, 15.14) | 0.009 |
| - De-escalation to more narrow-spectrum treatment | 10.3 (10/97) | 4.9 (5/103) | 5.5 (-1.9, 12.8) | 2.21 (0.74, 7.52) | 0.14 |
| - Initiated pathogen-directed antimicrobial treatment (without prior empirical antibiotic treatment) | 6.2 (6/97) | 0 (0/103) | 6.2 (1.4, 11.0) | NA | 0.01 |
| *Narrow-spectrum antibiotics within 48h* | 83.5 (81/97) | 84.5 (87/103) | -1.0 (-11.1, 9.2) | 0.93 (0.43, 1.99) | 0.85 |
| *Single dose of antibiotics only* | 4.3 (4/97) | 0 (0/103) | 4.3 (0.2, 8.4) | NA | 0.04 |
| *Antibiotics not used for more than 48h<sup>§§</sup>* | 14.4 (14/97) | 21.4 (22/103) | -6.9 (-17.5, 3.6) | 0.62 (0.29, 1.29) | 0.21 |
| *Treatment with intravenous antibiotics<sup>§§</sup>* | 68.0 (66/97) | 72.8 (75/103) | -4.8 (-17.4, 7.8) | 0.79 (0.43, 1.46) | 0.46 |
| **Outcomes on duration**<br>**(days)** |  |  |  | **Ratio of medians** |  |
| *Provision of any antibiotics during hospitalisation (n=191)* | 4.0 (2.9, 6.0)<br>(n=93) | 3.9 (2.1, 6.1)<br>(n=98) | 0.4 (-0.4, 1.1) | 1.04 (0.87, 1.25) | 0.63 |
| *Provision of intravenous antibiotics (n=178)* | 3.3 (2.6, 5.7)<br>(n=85) | 3.1 (2.1, 5.0)<br>(n=93) | 0.3 (-0.5, 1.0) | 1.08 (0.86, 1.34) | 0.51 |
| *Provision of broad-spectrum antibiotics (n=62)* | 3.8 (1.6, 5.9)<br>(n=37) | 3.9 (3.0, 8.8)<br>(n=25) | -1.3 (-2.9, 0.3) | 0.68 (0.42, 1.10) | 0.11 |
| *Time to administration of antibiotics (h) (n=191)* | 2.1 (1.3, 3.7)<br>(n=93) | 2.1 (1.1, 3.8)<br>(n=98) | 0.26 (-0.60, 1.12) | 1.14 (0.90, 1.44) | 0.55 |
| *Turnaround time (h)* | 4.0 (3.6, 4.5) | 68.2 (38.3, 95.0) | -53.8 (-48.7, -59.5) | 0.07 (0.06, 0.08) | <0.0001 |
|  |  |  |  | **Hazard ratio** |  |
| *Time to provision of pathogen-directed treatment (h)<sup>§</sup>* | 29.9 (25.9, 34.1) | 42.3 (39.5, 45.1) | -12.3 (-17.3, -7.3) | 3.45 (1.98, 6.02) | <0.0001 |

Data shown as percentage (count) for binary outcomes (on use), median (IQR) for continuous outcomes, and restricted mean and 95% confidence interval for the event-time outcome.
<sup>#</sup>: Differences were estimated as risk differences for binary outcomes (on use) and differences in medians for continuous outcomes (on duration) using unadjusted logistic and linear regression with logarithm-transformed outcomes, respectively. Odds ratios for binary outcomes (on use) and ratios of medians for continuous outcomes (on duration) were estimated using unadjusted logistic and linear regression with logarithm-transformed outcomes, respectively. p-values corresponding to testing ratios equal to 1 were reported except for one outcome (single dose of antibiotics) where it corresponds to testing the risk difference equal to 0.
<sup>§</sup>: Time elapsed from randomisation to provision of pathogen-directed treatment and within 48h.
<sup>§§</sup>: Within the first 7 days after study inclusion.
<sup>\*</sup>: p-value for testing risk difference equal to 0 was 0.03.
FAP-plus, BioFire FilmArray Pneumonia panel *plus* (bioMérieux S.A., Marcy-l'Etoile, France); CAP, community-acquired pneumonia; IQR, interquartile range; NA, not available.

The mean time to provision of pathogen-directed treatment was 29.9h in the FAP-plus arm and 42.3h in the standard-of-care arm (mean difference -12.3h, 95% CI -17.3– -7.3hh; p<0.0001) and a corresponding hazard ratio for FAP-plus compared to standard of care of 3.45 (95% CI 1.98–6.02). The turnaround time was significantly shorter for testing by FAP-plus compared to standard-of-care (difference 53.8h, 95% CI 48.7–59.5h; p<0.0001).

For the remaining secondary outcomes on the provision of narrow-spectrum antibiotics within 48h, antibiotics use for no more than 48h, treatment with intravenous antibiotics, and the duration of provision of antibiotics, intravenous antibiotics, and broad-spectrum antibiotics, no significant differences were found.

### Length of stay and clinical outcomes

Median LOS was 3.3 and 3.2 days for the FAP-plus and standard-of-care, respectively (difference 0.15 days, 95% CI -0.55–0.85 days; p=0.67; eTable 5 in Supplement 2). For clinical outcomes, 29 of 187 (16%) patients in the FAP-plus arm and 35 of 187 (19%) patients in the standard-of-care arm were readmitted (absolute risk difference -3.2%, 95% CI -10.8–4.4%; p=0.41), 9 (5%) and 7 (4%) patients died within 30 days (absolute risk difference 1.1%, 95% CI -3.0–5.2%; p=0.61), and 16 (9%) and 11 (6%) patients died within 90 days (absolute risk difference 2.7%, 95% CI -2.6–7.9%; p=0.32).

### Microbiological detections

The FAP-plus arm had a higher number of bacterial detections (175 vs. 72) and viral detections (74 vs. 63) compared to standard-of-care. When considering only the patients with confirmed CAP, the FAP-plus arm maintained a higher total number of bacterial (113 vs. 57) and viral detections (39 vs 33). The detailed breakdown of microbiological detections in the two arms is shown in eTable6 in Supplement 2.

### Adverse events

No serious adverse events were observed, and the number of adverse events was similar in both arms: for saline-induced sputum, seven were registered in the FAP-plus and eight in the standard-of-care arm (both arms: dyspnoea (6), non-severe tachycardia, rapidly resolved hypoxemia (5), nausea (1) and coughing (1), non-severe tachycardia (2). One patient experienced coughing during endotracheal aspiration in the FAP-plus arm.

## Discussion

This randomised controlled trial demonstrates that a rapid syndromic PCR pneumonia panel (FAP-plus) for LRT-pathogens as part of the diagnostic workup for hospitalised patients with suspected CAP increases the provision of and reduces the time to pathogen-directed treatment as compared to comprehensive standard-of-care microbiological testing. The FAP-plus panel provides clinicians in the emergency department with close to real-time information for actionable treatment decisions.

This is the first RCT on the effect of a comprehensive, rapid syndromic PCR pneumonia panel applied specifically to patients hospitalised with CAP. Most previous studies have not used a comprehensive syndromic PCR panel nor included patients shortly after admission, potentially limiting the advantages of rapid molecular testing. Another aspect of its novelty is the emphasis on pragmatism whereby decisions to continue, switch or discontinue antimicrobial treatment were solely at the treating physician’s discretion.

The FAP-plus led to a reduction in the mean-time without provision of pathogen-directed treatment within the first 48h from randomisation by approximately 10h compared to standard-of-care. Of note, for CAP patients, the median turnaround time (from admission to receiving an LRT-test result without restriction to 48h) was reduced much more (approximately 54h) for FAP-plus compared to standard-of-care. This result partly reflects hospital practice and is comparable to previous findings^12^ A faster microbiologic diagnosis allows for directed therapy, which has been shown to improve outcomes, limit antibiotic overuse, and prevent antimicrobial resistance^6,10^. Despite crowded conditions at the ED, a FAP-plus result was delivered within 4 hours for CAP patients, a turnaround time comparable to that achieved in other centres.^12^

While the study did not yield significant differences in clinical outcomes between the FAP-plus and standard-of-care groups, it’s important to note that the primary aim of this trial was to focus on diagnostic stewardship objectives by judiciously leveraging the rapid, multi-pathogen detection capabilities of the FAP-plus panel. We sought to reduce the time to provision of pathogen-directed treatment, potentially decreasing unnecessary or broad-spectrum antibiotic use and fostering antimicrobial stewardship. Future research should continue to explore innovative approaches to improving the diagnosis and management of respiratory infections, such as incorporating clinical decision support tools and antimicrobial stewardship programs into routine practice.

The strengths of this study include the pragmatic design, primary outcome values obtained from documentation in patient medical records by treating physicians not involved in the study, duration encompassing two winter seasons, the broad inclusion criteria representing typical patients with respiratory tract symptoms admitted to Norwegian hospitals, representative sampling from the LRT, a simple intervention, and comparison to comprehensive standard-of-care microbiological testing. These factors suggest that the primary findings of this study are likely generalizable to similar hospital settings. Standard-of-care in this study encompassed a wide battery of tests, including in-house molecular tests, rendering standard-of-care as competitive as possible in comparison with the commercial syndromic molecular panel. Moreover, the syndromic panel selected includes several bacterial and viral pathogens together with targets for selected antimicrobial resistance genes, making the panel applicable to other settings with different microbial etiological profiles and background resistance rates for CAP.^20^

A key strength derives from the fact that the study physicians were not involved in the treatment of patients. The restriction to a narrow time frame (48 h after randomisation), i.e., encompassing the period from respiratory sampling to the availability of results, supports interventions that can contribute to the timely administration of appropriate antibiotics, a central tenant of care for patients with pneumonia. The higher rate of continuation of appropriate empirical treatment for CAP patients in the FAP-plus arm suggests that this diagnostic tool assists in confirming the appropriateness of the initial empirical therapy and avoids unnecessary changes in treatment. Additionally, a larger proportion of patients in the FAP-plus arm received only a single dose of antibiotics, indicating the potential for reduced antibiotic exposure.

More patients in the FAP-plus arm than in the standard-of-care arm had an escalation from narrow-spectrum to more broad-spectrum antibiotics. This could raise concerns about antibiotic overuse; however, it is pertinent to emphasise that Norway has a low level of antibiotic resistance, and guidelines recommend the use of narrow-spectrum antibiotics ^21^. For the 200 CAP patients in this study, 141 (71.2%) received an initial empirical treatment that was in accordance with the national guidelines for antimicrobial treatment in hospitals (data not shown). As per guidelines, empirical treatment for mild to moderate CAP is benzylpenicillin and benzylpenicillin and gentamicin for severe CAP^21^. Treatment with narrow-spectrum antibiotics for RTIs is common practice in Norway,^22^ leaving little room for de-escalation, and even change from penicillin G to a more broad-spectrum penicillin (e.g., ampicillin) was considered an escalation. A background of a low level of antibiotic resistance implies that the differences found between the two arms for escalation or de-escalation of an antibiotic are, in a sense, the minimal differences to expect when introducing rapid testing in a Norwegian hospital setting. Of note, most escalation cases in our study involved a change from benzylpenicillin to more broad-spectrum penicillin, often ampicillin, or to a third-generation cephalosporin. It is important to consider that empirical treatment for CAP in many countries involves using a broad-spectrum beta-lactam antibiotic in combination with a macrolide. Changing to penicillin or third-generation cephalosporin is considered a de-escalation in this context.

Our study has some limitations. First, the single-centre design limits generalisability. However, our study demonstrates that embedding comprehensive rapid testing in a busy ED setting is possible. The trial was stopped early for efficacy, and there could be a risk of inflated estimates of differences between FAP-plus and standard-of-care, although this risk is presumably small as we found highly significant differences. Moreover, inflation is generally small, and continuing a trial to achieve a slight change in estimates would not be rational.^23^

In summary, rapid syndromic PCR testing for respiratory pathogens is associated with faster and more targeted microbial treatment for hospitalized patients with CAP. The study findings align with the broader concept of clinical management or treatment stewardship for lower respiratory tract infections. Routinely deployed rapid syndromic testing could complement or potentially replace targeted components of the standard laboratory-based diagnostic repertoire for patients admitted to the hospital with an acute respiratory illness. Future studies should focus on the impact of comprehensive rapid syndromic testing on clinical outcomes, the cost-effectiveness of this diagnostic approach, and the development of implementation strategies to facilitate the integration of rapid syndromic testing into routine clinical practice.

### Contributors

HMSG, CR, DFJ, and EU conceived and designed the study with advice from LH, TWC and CHVW. HMSG coordinated the overall study from inception to conclusion. EU and ROB were co-coordinators at HUH. SS and DLM recruited patients, performed patient assessments, collected data, or both. SS coordinated patient recruitment. SS and DLM compiled the data and DLM, SS, CR, and HMSG verified the data and take responsibility for the data’s integrity and the data analysis’s accuracy. CR analysed the data. ØK, MHE, STK, EU and HMSG coordinated the microbiology investigations. CR, SS, DLM, MHE, STK, and HMSG contributed to the methodology. CR, DLM, SS, and HMSG drafted the manuscript, and HMSG submitted the final version. All authors participated in the interpretation of the results and revised and approved the final version of the manuscript. All authors had final responsibility for the decision to submit for publication.

### Declaration of interests

TWC has received speaker fees, honoraria, travel reimbursement, and equipment and consumables at discount or free of charge for the purposes independent of research, outside of this submitted study, from BioFire diagnostics, BioMérieux and QIAGEN. TWC has received consultancy fees from Cepheid, Synairgen research, Randox laboratories and Cidara therapeutics. He has received honoraria for participation in advisory boards and consultancy fees from Cepheid, Roche, Janssen, and Shionogi. He is a member of the independent data monitoring committees for trials sponsored by Roche. He has acted as the UK chief investigator for studies sponsored by Janssen.

C. H. van Werkhoven has received financial research support from BioMérieux, DaVolterra, LimmaTech and consulting fees from Merck and Sanofi-Pasteur.

LH has received an honorarium as a scientific advisor in Pharmaholdings and holds shares in AlgiPharma.

All other authors have nothing to disclose.

## Data Availability

All de-identified and anonymized participant data analyzed and presented in this study are available from the corresponding author following publication upon reasonable request.
How to access data: Data sharing requests within three years of publication should be made to the corresponding author at.

## Acknowledgements

We thank all patients recruited at the Haukeland University Hospital (HUH), Bergen. We gratefully acknowledge the study nurses and staff at the Emergency Care Clinic (HUH), the study technologists and staff at the Department of Microbiology (HUH), the staff at the Biobank (HUH), the staff at the Section of Research and Innovation (HUH) and assistance from project administrators at the Department of Clinical Science, University of Bergen. The views expressed in this publication are those of the authors.

This work was supported financially by funding from the Research Council of Norway (NORCAP; 288718), the principal funder of the trial (ClinicalTrials.govNCT04660084). Co-funding was obtained from the Trond Mohn Foundation (COVID-19 CAPNOR; TMS2020TMT07 and RESPNOR; BFS2019TMT06), the University of Bergen, and Haukeland University Hospital. The funders had no role in the study design, collection, management, analysis, and interpretation of data, nor in the decision to submit this manuscript for publication.

## Notes

### Competing Interest Statement

The authors have declared no competing interest.

### Clinical Trial

NCT04660084

### Clinical Protocols

https://doi.org/10.1186/s13063-022-06467-7

### Author Declarations

The Regional Committee for Medical Research Ethics for South East Norway gave ethical approval for this work (case no. 31935).

