## Supplement for "Evaluating the Impact of Diagnostic Stewardship in Community-Acquired Pneumonia with Syndromic Molecular Testing: A Randomized Clinical Trial"

***Authors***

Dagfinn L. Markussen*, Sondre Serigstad*, Christian Ritz*, Siri T Knoop, Marit H Ebbesen, Daniel Faurholt-Jepsen, Lars Heggelund, C. H. (Henri) van Werkhoven, Tristan W Clark, Rune O Bjørneklett, Øyvind Kommedal, Elling Ulvestad and Harleen M.S. Grewal**

***Equal contribution***

**Department of Clinical Science, Bergen Integrated Diagnostic Stewardship cluster, University of Bergen, Bergen, Norway** (S Serigstad MD, DL Markussen MD, C Ritz PhD, ST Knoop MD, PhD, L Heggelund MD, PhD, Elling Ulvestad MD, PhD, HMS Grewal MD, PhD) **and Emergency Care Clinic, Haukeland University Hospital, Bergen, Norway** (S Serigstad MD, DL Markussen MD, R Bjørneklett MD, PhD) **and Department of Clinical Medicine, University of Bergen, Bergen, Norway** (S Serigstad MD, R Bjørneklett MD, PhD) and National Institute of Public Health, University of Southern Denmark, Copenhagen, Denmark (C Ritz PhD) and Department of Microbiology, Haukeland University Hospital, Bergen, Norway (ST Knoop MD, PhD, MH Ebbesen MD, PhD, Ø Kommedal MD, PhD, E Ulvestad MD, PhD, HMS Grewal MD, PhD) **and Department of Infectious Diseases, Rigshospitalet, Denmark** (D Faurholt-Jepsen MD, PhD) **and Department of Internal Medicine, Vestre Viken Hospital Trust, Drammen, Norway** (L Heggelund MD, PhD) **and Julius Center for Health Sciences and Primary Care, University Medical Center Utrecht, Utrecht, The Netherlands** (C. H. van Werkhoven MD, PhD**) and School of Clinical and Experimental Sciences, Faculty of Medicine, University of Southampton, Southampton, UK** (TW Clark MD)

**Table of contents**

### Contents

| | | |
|---|---|---|
| eTable 1 | Standard comments for detected bacteria. | 3 |
| References | | 4 |
| eTable 2 | Definitions and diagnostic clinical criteria for CAP. | 5 |
| References | | 5 |
| eTable 3 | Baseline characteristics for CAP patients. | 6 |
| 4 | Pathogen-directed treatment within 48 hours for patients with pneumonia. | 8 |
| eTable 5 | Comparison of length of stay and clinical outcomes between rapid testing by FAP *plus* and standard-of-care for all randomised patients and for CAP patients only. | 9 |
| eTable 6 | Microbiological detections in respiratory samples in the standard-of-care and FAP *plus* arms. | 10 |

**eTable 1.**

**Standard comments for detected bacteria [3-20]**

| Respiratory tract microbes divided in different categories according to their potential clinical relevance | |
|---|---|
| **Category** | **Microbe** |
| **A: Always pathogens**<br>Always considered relevant in a patient with respiratory tract infection | Adenovirus [6,8,12,14]<br>Coronavirus [8,12,14]<br>Human metapneumovirus [6, 12,14]<br>Influenza virus [6,8,12,14]<br>MERS [9]<br>Parainfluenza virus [8,12,14]<br>Rhinovirus [6,12,14]<br>RS-virus [6,8,12,14]<br><br>*Bordetella pertussis* [4]<br>*Chlamydia pneumoniae* [3,6,8,12,14]<br>*Legionella pneumophila* [3,6,14]<br>*Mycoplasma pneumoniae* [3,6,8,12,14] |
| **B: Usually pathogens**<br>Can be colonizers, but are usually considered relevant in a patient with pneumonia | *Haemophilus influenzae* [3,6,8,12,14]<br>*Streptococcus pneumoniae* [3,6,8,12,14,20]<br>*Streptococcus pyogenes* [11,18,20] |
| **C: Usually not pathogens**<br>Usually colonizers, but can cause pneumonia, especially in patients with chronic diseases | *Klebsiella pneumoniae* [7,14,20]<br>*Moraxella catarrhalis* [3,5,12,14]<br>*Staphylococcus aureus* [3,12,14,15,20] |
| **D: Usually not pathogens**<br>A seldom cause of pneumonia in otherwise healthy patients. May be considered relevant if detected as the only bacterial species in a patient with immunosuppression and/or several previous courses of antibiotics | *Acinetobacter calcoaceticus-baumannii-complex* [4,12,20]<br>*Enterobacter cloacae* [6,12,19]<br>*Escherichia coli* [6,12,14,19,20]<br>*Klebsiella aerogenes* [6,12]<br>*Klebsiella oxytoca* [6,12]<br>*Proteus species* [6,12,19]<br>*Pseudomonas aeruginosa* [12-14,19,20]<br>*Serratia marcescens* [6,12,19]<br>*Streptococcus agalactiae* [17] |

#### eTable 2

**Definitions and diagnostic clinical criteria for CAP** (Adapted from Postma D.F. et al. [1] and Serigstad S *et al* [2])

Clinical CAP:

- Patients with at least two diagnostic criteria* and in-hospital treatment and/or diagnosis of clinically suspected CAP documented by the treating physician and in agreement with the assessment by a study investigator. In case of disagreement, an additional study investigator will arbitrate.
- Patients with two or more diagnostic criteria and an obvious non-respiratory source of infection are not considered to have a clinical diagnosis of CAP.

Radiologically confirmed CAP:

- Clinical CAP
- The presence of a new or increased infiltrate on chest radiography or computer tomography (CT).

*** Diagnostic criteria:**

- Cough: recent or worsening
- Production of purulent sputum or a change in the character of sputum
- Temperature >38.0°C or <36.1°C
- Auscultatory findings consistent with pneumonia, including rales, evidence of pulmonary consolidation (dullness on percussion, bronchial breath sounds, or egophony), or both
- Leukocytosis (>11.0 × $10^9$ white cells per liter or > 8.2 x $10^9$ neutrophils)
- C-reactive protein (CRP) level > 50 mg/L
- Dyspnea, tachypnea, or hypoxemia: recent or worsening

#### eTable 3

**Baseline characteristics for CAP patients only (n=200)**

| | **FAP *plus*** (n=97) | **Standard-of-care** (n=103) |
|---|---|---|
| *Age (years)* | 71 (60, 82) | 72 (62, 80) |
| *Sex (% Female)* | 40 (39/97) | 42 (43/103) |
| *Current smoker* | 21 (20/97) | 13 (14/103) |
| *Influenza vaccine* | 53 (52/97) | 58 (60/103) |
| *Pneumococcal vaccine* | 38 (37/97) | 37 (38/103) |
| *Duration of symptoms (days)* | 6.5 (3.6, 8.7) | 5.8 (3.5, 7.6) |
| *Antibiotics with 48 hours* | 23 (22/97) | 17 (18/103) |
| *Antibiotics within the last month (%)* | 15 (15/97) | 9 (9/103) |
| ***Comorbidities*** | | |
| *Hypertension* | 37 (36/97) | 39 (40/103) |
| *Cardiovascular disease* | 34 (33/97) | 30 (31/103) |
| *Respiratory disease* | 54 (52/97) | 50 (51/103) |
| *Renal disease* | 11 (11/97) | 9 (9/103) |
| *Liver disease* | 0 (0/97) | 2 (2/103) |
| *Diabetes* | 8 (8/97) | 14 (14/103) |
| *Immunocompromised* | 8 (8/97) | 12 (12/103) |
| *Cancer* | 9 (9/97) | 7 (7/103) |
| *Charlson Comorbidity Index score* | 4 (2, 5) | 4 (2, 5) |
| ***Observations*** | | |
| *Temperature (°C)* | 37.2 (36.8, 37.6) | 37.2 (36.8, 37.7) |
| *Pulse rate (bpm)* | 92 (78, 109) | 94 (85, 110) |

| | | |
|---|---|---|
| *Respiratory rate (bpm)* | 22 (20, 25) | 24 (20, 28) |
| *$O_2$ saturations (%)* | 93 (90, 96) | 93 (89, 96) |
| *Supplementary $O_2$* | 6 (6/97) | 6 (6/103) |
| ***Blood pressure (mmHg)*** | | |
| *Systolic* | 127 (116, 146) | 134 (117, 148) |
| *Diastolic* | 80 (71, 86) | 80 (70, 88) |
| ***Laboratory and radiology*** | | |
| *CRP (mg/L)* | 135 (83, 209) | 151 (101, 237) |
| *White blood cell count ($\times 10^9$ per L)* | 11.3 (8.1, 16.7) | 11.9 (8.7, 15.0) |
| *Chest X-ray (%)* | 96 (93/97) | 97 (100/103) |
| *Chest CT (%)* | 24 (23/97) | 20 (21/103) |
| *CRB-65* | 1 (1, 2) | 1 (1, 2) |
| *qSOFA* | 1 (0, 1) | 1 (0, 1) |
| *SARS-CoV-2 positive test* | 15 (15/97) | 16 (16/103) |

Data are shown as percentage (count) and median (IQR) for binary and continuous variables, respectively.

IQR = interquartile range. $O_2$=oxygen. CRP = C-reactive protein. CT = computed tomography. CRP and white blood cell counts presented are the highest values during hospitalization. *CRB-65=* confusion, respiratory rate, blood pressure, and age > 65 years; qSOFA= quick sequential organ failure assessment score.

#### eTable 4

**Pathogen-directed treatment within 48 hours for patients with pneumonia (n=200)**

| **Pathogen-directed treatment** | **Test that initiated pathogen-directed treatment** | **Standard of care N=16** | | **FAP *plus* N=46** | |
|---|---|---|---|---|---|
| | | N | Pathogen-directed antibacterial therapy | N | Pathogen-directed antibacterial therapy |
| **Continuation of appropriate empirical treatment** | Blood cultures | 0 | | 2 | PenG: 2 |
| | LRT culture | 1 | PenG: 1 | 0 | |
| | FAP *plus* | 0 | | 13 | PenG: 11<br>Carbapenems: 1<br>Pip/Tazo: 1 |
| | PUAT | 6 | PenG: 5<br>Ampicillin: 1 | 1 | PenG: 1 |
| | Total | 7 | | 16 | |
| **Escalation to more broad-spectrum treatment** | Blood cultures | 3 | PenG to 3GC: 2<br>PenG to a more broad-spectrum penicillin: 1 | 0 | |
| | LRT culture | 1 | PenG to a more broad-spectrum penicillin: 1 | 0 | |
| | FAP *plus* | 0 | | 14 | PenG changed to a more broad-spectrum penicillin: 6<br>Benzylpenicillin to 3GC: 5<br>Aminoglycoside added: 2<br>Other: 1 |
| | Total | 4 | | 14 | |
| **De-escalation to more narrow-spectrum treatment** | Other | 1* | 3GC to quinolone | 0 | |
| | LRT culture | 2 | | 0 | |
| | FAP *plus* | 0 | | 10 | Stopped aminoglycosides: 5<br>Stopped macrolides: 3<br>Stopped clindamycin: 1<br>3GC to ciprofloxacin: 1 |
| | PUAT | 4 | Stopped aminoglycosides: 3<br>3GC to PenG: 1 | 0 | |
| | Total | 5 | | 10 | |
| **Initiated pathogen-directed antimicrobial treatment (without prior empirical antibiotic treatment)** | FAP *plus* | 0 | | 6 | PenG: 2<br>Ampicillin: 1<br>Amoxi/Clav:1<br>3GC: 2 |
| | Total | 0 | | 6 | |

Abbreviations: FAP *plus* - Biofire® Filmarray® Pneumonia plus Panel; LRT culture – lower respiratory tract sample culture; PUAT – pneumococcal urine antigen test; PenG – Benzylpenicillin; 3GC - third generation cephalosporins; Pip/Tazo - Piperacillin/Tazobactam; TMP/S - Trimethoprim/sulfamethoxazole; Amoxi/Clav - Amoxicillin/Clavulanic acid

*) *Legionella pneumohilia* PCR

#### eTable 5

**Comparison of length of stay and clinical outcomes between rapid testing by FAP *plus* and standard-of-care for all randomised patients (n=374) and CAP patients only (n=200), respectively**

| | FAP *plus* | Standard-of-care | FAP *plus* vs. Standard-of-care# | | |
|---|---|---|---|---|---|
| ***All patients** (n=374)* | | | **Difference** | **Odds ratio** | **p-value** |
| *Readmission* | 15.5 (29/187) | 18.7 (35/187) | -3.2 (-10.8, 4.4) | 0.80 (0.46, 1.37) | 0.41 |
| *30-day mortality* | 4.8 (9/187) | 3.7 (7/187) | 1.1 (-3.0, 5.2) | 1.30 (0.47, 3.71) | 0.61 |
| *90-day mortality* | 8.6 (16/187) | 5.9 (11/187) | 2.7 (-2.6, 7.9) | 1.50 (0.68, 3.41) | 0.32 |
| | | | | **Ratio of medians** | |
| *Length of stay (days)* | 3.3 (2.0, 6.0) | 3.2 (2.0, 6.0) | 0.15 (-0.55, 0.85) | 1.05 (0.83, 1.34) | 0.67 |
| ***CAP patients** (n=200)* | | | | **Odds ratio** | |
| *Readmission* | 14.4 (14/97) | 18.4 (19/103) | -4.0 (-14.3, 6.2) | 0.75 (0.35, 1.58) | 0.45 |
| *30-day mortality* | 3.1 (3/97) | 3.9 (4/103) | -0.8 (-5.9, 4.3) | 0.79 (0.15, 3.67) | 0.76 |
| *90-day mortality* | 7.2 (7/97) | 5.8 (6/103) | 1.4 (-5.5, 8.2) | 1.26 (0.40, 4.04) | 0.69 |
| | | | | **Ratio of medians** | |
| *Length of stay (days)* | 3.9 (2.9, 6.1) | 4.0 (2.2, 6.3) | 0.48 (-0.48, 1.45) | 1.15 (0.87, 1.51) | 0.33 |

Data shown as percentage (count) for binary outcomes and as median (IQR) for length of stay.

#: Differences were estimated as risk differences for binary outcomes (on readmission and mortality) and differences of median for length of stay using unadjusted logistic regression and linear regression with logarithm-transformed length of stay, respectively. Odds ratios for binary outcomes and ratio of medians for length of stay were estimated using unadjusted logistic and linear regression with logarithm-transformed length of stay, respectively. p-values corresponding to testing ratios equal to 1 were reported.

FAP *plus*, BioFire FilmArray Pneumonia panel *plus* (bioMérieux S.A., Marcy-l'Etoile, France); CAP, community-acquired pneumonia; IQR, interquartile range.

#### eTable 6

**Microbiological detections in respiratory samples in the standard-of-care and FAP *plus* arms**

| Microbe detected | All randomized patients | | Patients with confirmed pneumonia | |
|---|---|---|---|---|
| | Standard of care (n=187) | FAP plus (n=187) | Standard of care (n=106) | FAP plus (n=102) |
| *Acinetobacter calcoaceticus-baumannii* complex | 0 | 4 | 0 | 1 |
| *Enterobacter cloacae* complex | 0 | 4 | 0 | 4 |
| *Escherichia coli* | 5 | 13 | 3 | 5 |
| *Haemophilus influenzae* | 22 | 40 | 18 | 29 |
| *Klebsiella aerogenes* | 0 | 0 | 0 | 0 |
| *Klebsiella oxytoca* | 1 | 6 | 1 | 3 |
| *Klebsiella pneumoniae* group | 2 | 5 | 1 | 3 |
| *Moraxella catarrhalis* | 11 | 16 | 9 | 11 |
| *Proteus* spp. | 0 | 3 | 0 | 1 |
| *Pseudomonas aeruginosa* | 5 | 10 | 4 | 4 |
| *Serratia marcescens* | 2 | 2 | 0 | 1 |
| *Staphylococcus aureus* | 7 | 36 | 4 | 23 |
| *Streptococcus agalactiae* | 0 | 9 | 0 | 6 |
| *Streptococcus pneumoniae* | 14 | 26 | 14 | 21 |
| *Streptococcus pyogenes* | 0 | 1 | 0 | 1 |
| *Legionella pneumophila* | 1 | 0 | 1 | 0 |
| *Mycoplasma pneumoniae* | 0 | 0 | 0 | 0 |
| *Chlamydia pneumoniae* | 0 | 0 | 0 | 0 |
| Other bacterial pathogens not included in the FAP *plus* panel* | 3 | 0 | 2 | 0 |
| **Total number of bacterial detections** | **72** | **175** | **57** | **113** |
| SARS-CoV-2** | 24 | 28 | 17 | 16 |
| Human rhinovirus/enterovirus | 14 | 22 | 5 | 13 |
| Parainfluenza virus | 7 | 4 | 2 | 0 |
| Respiratory syncytial virus | 10 | 15 | 4 | 7 |
| Seasonal coronavirus*** | 0 | 7 | 0 | 5 |
| Human metapneumovirus | 7 | 0 | 5 | 0 |
| Adenovirus | 0 | 1 | 0 | 0 |
| Influenza A virus | 1 | 0 | 1 | 0 |
| Influenza B virus | 1 | 0 | 1 | 0 |
| **Total number of viral detections** | **63** | **74** | **33** | **39** |

Abbreviations: FAP *plus*, Biofire FilmArray Pneumonia panel *plus*; SARS-CoV-2, severe acute respiratory syndrome coronavirus 2.

*) One detection of *S. dysgalactiae* and two detections of *K. variicola*

**) SARS-CoV-2 is not a target in the FAP *plus* and was tested using laboratory PCR.

***) Seasonal human coronavirus includes: HKU1, 229E, NL63 and OC43. Testing for seasonal coronavirus was not part of the standard of care diagnostics until May 2022.
